# Assessment of risk factors associated with and practices of cattle farmers in Kirehe District Rwanda with respect to vector-borne and zoonotic pathogens

**DOI:** 10.1101/2024.05.04.24306778

**Authors:** M. Fausta Dutuze, Analise Espino, Rebecca C. Christofferson

## Abstract

Rural Rwandan communities face challenges to the health of humans and animals. The topography and climate of the Kirehe District of Rwanda put farmers there at high risk for mosquito-borne diseases. Farmers from 92 Rwandan farms were surveyed about farm practices, as well as animal and human health histories between December 2017 to February 2018. Further, human, animal, and environmental factors were investigated to determine whether there is a pattern in risk for abortion occurrence in cattle and/or history of malarial disease in the human populations. Iterative, complimentary logistic regression models were employed to determine whether there was an association between surveyed variables and spontaneous cattle abortion, a known symptom of and risk factor for Rift Valley Fever virus and other mosquito-borne/zoonotic pathogens in the region. These factors were then used to investigate association with a reported history of malaria. Of the 92 farms in our study, 82 were family farms and 10 were commercial farms. On average, 88% of the farms had cattle and 30% of farms had experienced a cattle abortion in the past two years. There was no discernable pattern in significance in the risk factors for history of abortion in cattle. This One Health approach sought to investigate human, animal, and environmental factors surrounding the transmission of zoonotic, mosquito-borne viruses. From our study into the practices of the farmers with respect to biosafety and self-protection against disease, we identify potential sources of risk that could be targeted for enhance education and protection on these farms.

## Introduction

Vector-borne diseases account for 17% of all infectious diseases and 700,000 deaths annually [1, 2]. Rift Valley fever virus (RVFV) is a mosquito-borne viral zoonosis, a disease naturally transmissible between animals and people [3]. This virus was first isolated in 1931 in Kenya’s Great Rift Valley region when there were 4,700 lamb deaths in four weeks [3]. In 2000, 70 years after RVFV’s isolation, the first outbreak outside of Africa was reported and RVFV has since become a global priority disease [4]. Heavy rainfall usually precedes RVFV outbreaks, as it is spread by the *Aedes* and *Culex* mosquitoes [4]. While RVFV in humans is usually mild, a few cases progress to a severe form of the disease [4, 5]. It is estimated that up to 2% of RVFV human cases result in ocular disease, <1% result in meningoencephalitis, and <1% develop hemorrhagic fever [4, 5]. Overall, the human cases fatality rate is <1%, with the majority of fatalities occurring in patients who developed hemorrhagic fever [4, 5]. However, the mortality rate of RVFV in all animals is much higher, with the mortality rate of sheep and lambs together at 20-70% and that of young lambs alone as high as 70-100% [4, 5]. Furthermore, the rate of spontaneous abortion in pregnant sheep and cattle with RVFV is 80-100% [4, 5].

Given that RVFV significantly affects cattle and is listed as a priority pathogen by the World Health Organization, understanding the eco-epidemiological factors that describe the One Health landscape of these communities is essential for assessing risk [6, 7]. Part of that understanding includes the landscape of RVFV clinical presentation, including abortions, to clarify the true burden of RVFV and identify potential alternative etiologies.

The Republic of Rwanda is in the Great Rift Valley of Central Africa and is known as “the land of a thousand hills,” contributing to its temperature and humidity variability. As of 2021, Rwanda is ranked second highest in population density with one of the smallest African countries by area but with a population of 13.46 million [8–10]. The UN World Population Prospects data from 2022 shows that the Rwandan population has increased by nearly 2.5% yearly since 2004 [11]. While an increase in urbanization has worked in tandem with this increase in population, the country still has an 80% rural population [10]. Rural communities are known to have poorer health outcomes due to inequalities in access to healthcare and health coverage. This medical inequality has been associated with lower poverty and educational status [12]. The EICV5 Main Indicators Report shows that 53% of workers identify as independent farmers, a category with a 34% working poverty rate [13].

Rwanda is divided into five provinces—Northern, Southern, Western, Eastern, and Kigali—which can be further subdivided into sectors, cells, and villages. The Eastern Province is a prime location for mosquitos, given the low altitudes, precipitation, and proximity to lakes and plantations. A study in 2011 on the most prevalent mosquito species in this area found four species—*Culex* spp., *Aedes* spp., *Coquillettidia* spp., and *Anopheles* spp.—that are known to be primary and secondary vectors of Rift Valley Fever virus and malaria [14]. A study completed in the western region of Uganda, which has a similar landscape to the Eastern Province of Rwanda, found a 10.4% overall RVFV seroprevalence [15]. Of all the livestock, cattle had nearly double RVFV seroprevalence compared to other livestock (20.5%); sheep and goats followed with 6.8% and 3.6% seroprevalences, respectively [15].

Our study was conducted in the Eastern Province. According to the Fifth Integrated Living Conditions Survey (EICV5) Poverty Profile Report, in 2016, 37.4% of the population in the Eastern Province lived in poverty, and 15.3% lived in extreme poverty [16]. Rwanda’s Fourth Integrated Household Living Conditions Survey from 2013-2014 showed that nearly 70% of Rwandans in the working population are employed in agriculture, and in the Eastern Province of Rwanda, 67% of households raise livestock [17]. Nationally, there has been an increase of three percentage points in the percentage of households raising cattle from 2010-2013 [17].

Rural Rwandan families are the target demographic for this program, as families without cattle are associated with a lower socioeconomic class [18, 19]. The Eastern Province has the highest participation of Girinka at a rate of 10% [17]. Between 2006 and 2017, the program distributed nearly 300,000 cattle and helped over one million individuals [20]. The Girinka report from 2014 highlighted high citizen satisfaction in the following roles of the program: fighting malnutrition (97%), fighting poverty (96%), and promoting social cohesion (95%) [20]. A controlled cohort study in Rwanda’s Nyabihu and Ruhango districts found that the Girinka program was strongly associated with reduced stunting in children [21]. Another study in the Bugesera district in 2013 found that the Girinka program was essential in increasing food security and eliminating child malnutrition [22]. Beyond its health benefits, Girinka has successfully increased agricultural production and, ultimately, increased the social class of the beneficiaries [21, 22]. Kim *et al*. found that over 90% of beneficiaries used their manure and attributed their increased crop yields to better soil fertility [23]. Manure is known to help crop production and is associated with increased food security [24]. Overall, the Girinka program has been perceived by citizens as successful and has been associated with alleviating health and financial hardship.

However, Rwanda experienced its largest RVFV outbreak in 2018 following unusually extreme rainfall [14]. This outbreak also affected other regions of eastern Africa, such as Kenya, Uganda, and Tanzania, before ending in July 2018 [14]. During this outbreak, many livestock deaths and abortions were reported [14]. While other etiologies were identified, many cases of cattle abortion and all cases of cattle hemorrhagic fever were confirmed to be PCR-positive for RVFV. Proximity to cattle is a risk factor for exposure to RVFV, yet the attitudes and practices of Rwandan farmers for basic knowledge of biosafety/biosecurity practices for this and associated pathogens remain understudied.

One Health encourages a holistic approach to determinants of health in the framework of the interaction of environmental, animal, and human health. The WHO describes One Health as involving “public health, veterinary, and environmental sectors [7].” The One Health approach is particularly relevant for food and water safety, nutrition, and controlling zoonoses. These diseases, such as flu, rabies, and Rift Valley fever, can spread between animals and humans[2, 25]. Further, systems thinking promotes thinking beyond the target question to determine if other elements in the interconnected system could be exploited or pursued to find holistic solutions to health issues.

## Methodology

In-person surveys were administered between December 2017 and February 2018 in the Kirehe District, Eastern Province (*Iburasirazuba*) of Rwanda, with the appropriate ethics approvals from the University of Rwanda, College of Medicine and Health Sciences (No. 377/CMHS IRB/2017). The enumerators conducted the in-person questionnaires in Kinyarwanda and then translated them into English for data analysis. The survey instrument can be found in **Supplemental Information S2**. A cluster-based method for data sampling was utilized, resulting in a total of 92 farms. The farm data was collected from the following sectors in the Kirehe district: Gahara (11), Kigarama (10), Kirehe (9), Mahama (7), Mpanga (10), Nasho (9), Nyamugari(19), & Nyarubuye (17) (**Figure 1**). Measurements were taken and recorded by study personnel to judge distance to points of interest. Study personnel also directly observed water collection and collection device types. Altitude, temperature, and humidity were measured using a handheld GPS with Altimeter and Compass (Garmin Ltd.)

**Figure 1:**
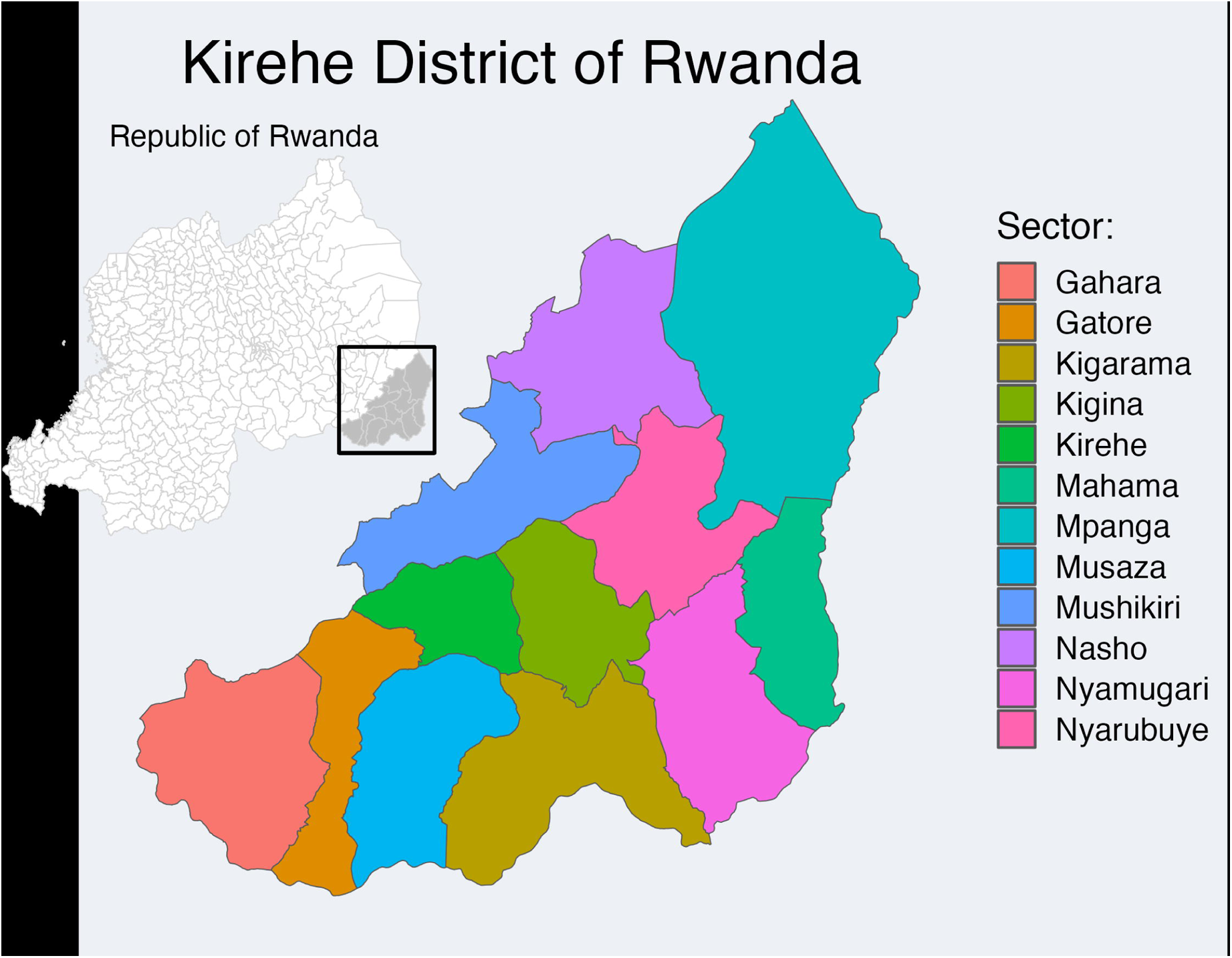
Rwanda (inset) with the Kirehe District highlighted and the sampled sectors color coded.

Variable Classification: We classified the following variables as environmental variables: farm type; altitude; temperature; humidity; garbage disposal; presence and distance to bush, swamp, and/or banana plants; and study personnel observed water retention in exposed containers. (Note: all farms had experienced rain within three days of site visits.) The following were classified as animal variables: number and source of cattle; history of cattle on the farm; presence of other animal species and housing; isolation of new cattle; and zero grazing practices. The following were classified as human variables: distance to human housing of cattle, bed net usage, history of malaria, and source and storage of water habits.

Univariate analysis was completed to examine whether there were differences in farm humidity, temperature, and altitude across Kirehe sectors. Logistic regression models were employed to examine any association between a history of spontaneous abortion in cattle on the farm with the variables of interest for each One Health vertex a) environmental factors, b) animal health, and c) human. Variables determined significant in each vertex were then put into a single model to interrogate associations with both abortion history in cattle and malaria history in humans. RStudio version 2023.06.1 was used for all analyses.

## Results

### Sectors of Eastern Province, Rwanda

There were some differences in the environmental characteristics across sectors in the Kirehe District. The average altitude of all sites included was 1462.6 meters above sea level. Using Kruskal-Wallis, it was determined that altitude was significantly different across sectors (p <0.0001). Subsequent pairwise comparisons (Wilcoxon rank sum test) show where differences lie (**Table 1**). The altitude means, minimums, and maximums by sector are in **Figure S1A**.

**Table 1.** Mean altitude (in meters) of each sector and range. *Indicates significance at the α=0.05 based on pairwise comparisons of altitude by sector using Wilcoxon rank sum test with Bonferroni p-value adjustment method.

|  | **Gahara**<br>**(1511**<br>**m)** | **Kigarama**<br>**(1676 m)** | **Kirehe**<br>**(1319**<br>**m)** | **Mahama**<br>**(1362 m)** | **Mpanga**<br>**(1322**<br>**m)** | **Nasho**<br>**(1345**<br>**m)** | **Nyamugari**<br>**(1371 m)** |
| --- | --- | --- | --- | --- | --- | --- | --- |
| **Kigarama** | 0.0023* | --- | --- | --- | --- | --- | --- |
| **Kirehe** | 0.2754 | 0.0041* | --- | --- | --- | --- | --- |
| **Mahama** | 0.0068* | 0.0086* | 1.000 | --- | --- | --- | --- |
| **Mpanga** | 0.0023* | 0.0031* | 1.000 | 0.0560 | --- | --- | --- |
| **Nasho** | 0.0031* | 0.0041* | 1.000 | 0.0182* | 0.0368* | --- | --- |
| **Nyamugari** | 0.0002* | 0.0004* | 1.000 | 1.000 | 0.0022* | 0.0385* | --- |
| **Nyarubuye**<br>**(1671 m)** | 0.0003* | 0.2828 | 0.001* | 0.0031* | 0.0005* | 0.0001* | <.0001* |

The overall average temperature was 24°C across all sites, but again, the temperature was significantly different among sectors (p <0.0001) and subsequently determined pairwise differences (**Table 2**). The temperature means, minimums, and maximums by sector are in **Figure S1B**.

**Table 2.** Table of pairwise comparisons of temperature by sector using Wilcoxon rank sum test with Bonferroni p-value adjustment method. *Indicates significance at the α=0.05.

|  | **Gahara** | **Kigarama** | **Kirehe** | **Mahama** | **Mpanga** | **Nasho** | **Nyamugari** |
| --- | --- | --- | --- | --- | --- | --- | --- |
| **Kigarama** | 0.0304* | --- | --- | --- | --- | --- | --- |
| **Kirehe** | 0.6897 | 1.000 | --- | --- | --- | --- | --- |
| **Mahama** | 0.0114* | 0.1516 | 0.1158 | --- | --- | --- | --- |
| **Mpanga** | 1.000 | 0.0049* | 0.8426 | 0.0177* | --- | --- | --- |
| **Nasho** | 0.2927 | 1.000 | 1.000 | 1.000 | 0.0598 | --- | --- |
| **Nyamugari** | 0.0058* | 1.000 | 1.000 | 1.000 | 0.0010* | 0.9879 | --- |
| **Nyarubuye** | 0.0019* | 1.000 | 0.5017 | 1.000 | 0.00010* | 0.8426 | 1.000 |

The average humidity of the farms was 64%, and again, we found a significant difference among sectors (p <0.0001). **Table 3** shows the pairwise comparison of humidity across sectors. **Figure S1C** shows the humidity minimum, maximum, and averages by sector.

**Table 3.**
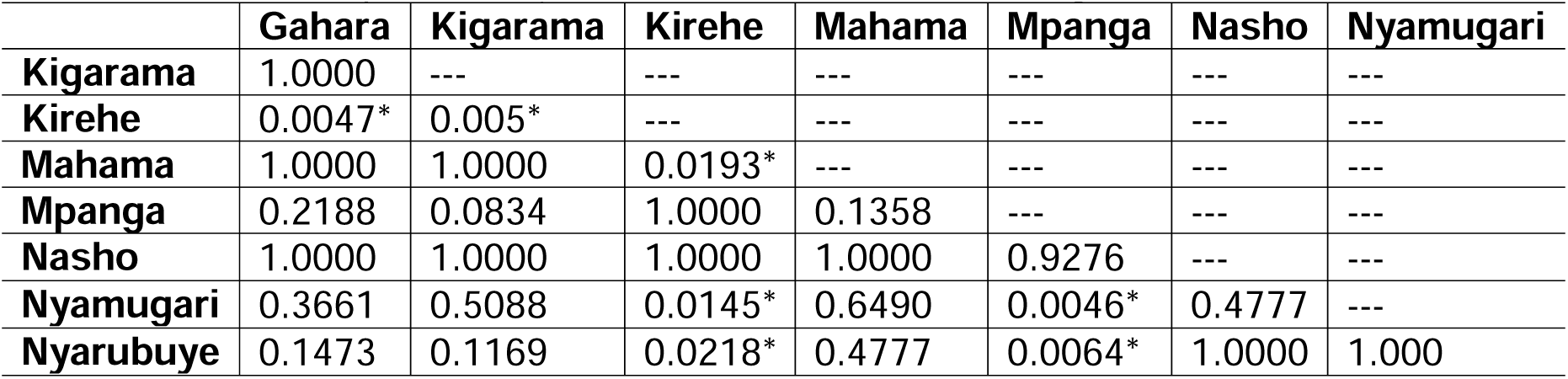
Table of pairwise comparisons of humidity by sector using Wilcoxon rank sum test with Bonferroni p-value adjustment method. *Indicates significance at the α=0.05.

Across all sectors, temperature was highly predictive of humidity (%) with a slope of - 3.26 and an Adjusted R^2^ of 0.6167 (p <0.0001, **Figure 2**). Altitude was only moderately associated with temperature and humidity with slopes -0.003382 and 0.020863, and adjusted R^2^ of 0.04628 and 0.1077, respectively (**Supplemental Figure S2**).

**Figure 2:**
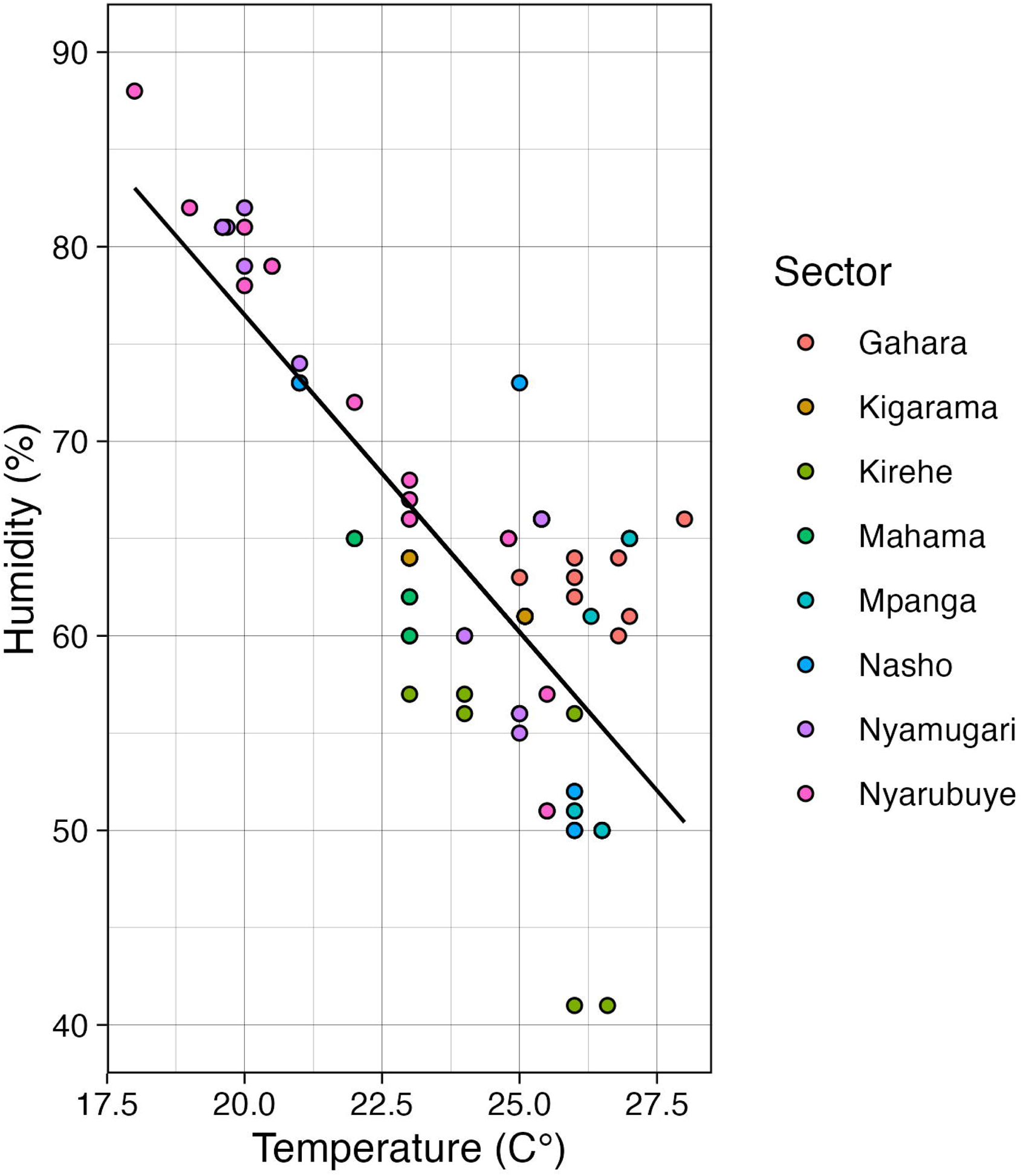
Linear association between temperature and humidity was statistically significantly (p<0.0001).

### Farm Demographics

A total of 92 farms were surveyed. The majority (90%) of the farms in our study were family farms (n = 82, 89.13%), and only ten were commercial farms (10.87%). Most farms had cows (90% and 87.8% of commercial and family farms, respectively), and many had small ruminants, namely, sheep or goats (40% and 62.41% of commercial and family farms, respectively). Few farms had poultry, only 13.4% of family farms (**Table 4**). Of those farms with cattle, 77.78% (n=7) of commercial and 34.15% of family farms had non-indigenous cattle breeds, while 44.44% of commercial and 47.56% of the family had local breeds. Interestingly, only 3 (3.66%) family farms reported a mix of exotic and local cattle breeds.

**Table 4:** Farm Demographics. . Numbers refer to the number of respondent farms that indicated “yes” to the question.

|  | Family<br>(n=82) | Commercial<br>(n=10) |
| --- | --- | --- |
| **Animal Makeup** |  |  |
| Cows | 72 | 9 |
| Small ruminants | 4 | 52 |
| Poultry | 0 | 11 |
| **Animal Housing** |  |  |
| Species pen separation | 56 | 7 |
| Shared pens | 21 | 1 |
| No housing | 5 | 2 |
| **Animal Practices** |  |  |
| Zero Grazing | 49 | 4 |
| Cattle from Government | 22 | 4 |
| Artificial Insemination (AI) | 7 | 5 |
| Natural Insemination (NI) | 68 | 4 |
| Both AI and NI | 6 | 1 |
| **Biosecurity** |  |  |
| Foot Bath | 0 | 0 |
| Isolation of new animals | 0 | 0 |
| Mosquito Control Practices – Human | 81 | 5 |
| Mosquito Control Practices – Animal | 2 | 3 |
| Abortive Tissue Disposal With Disinfectant | 0 | 0 |

Most farms (56 and 7 of family and commercial, respectively) practiced species separation with respect to housing, while the remaining had either no separation or no housing (**Table 4**). Zero grazing was practiced by 59.76% of family farms and 40% of commercial farms. Of those family farms with cattle, 30.56% (n = 22) indicated the government had been the source of the animal. The remaining farms received their cattle from family, dowry, neighbors, of the market. Of the commercial farms with cattle, 4 indicated the source was the government, with the remaining coming from the market (n = 4) or family (n = 2). Most cattle were bred using natural insemination, though some artificial insemination was practiced, with public veterinary officers exclusively identified as the source of semen. For natural insemination across both farm types, most (n = 75) males were sourced from neighbors.

On average, family farms had approximately three cattle, while commercial farms had approximately 26 cattle. Commercial farms reported an average of 20 exotic cattle less than one on average exotic for family farms. Commercial farms reported an average of 5 indigenous breeds of cattle per farm, while family farms reported approximately 2 indigenous cattle per farm on average (**Supplemental Table S1**).

All commercial and most (70.73%) of family farms reported being near the bush, with a median distance measured of 16 meters (IQR: 7-43.8m) for commercial and 7.5 meters (IQR: 3.75-50 meters) for family farms (**Supplemental Figure S3**). The distance to the bush was not significantly different between farm types (p = 0.4844). Half of the commercial farms reported being near swamps, while 36.59% of family farms reported being near swamps. However, the distance (in meters) to the nearest swamp was measured for all but three farms (**Supplemental Figure S4)**.

The commercial farms were closer to the swamps, with an average distance of 625 meters, while family farms were an average of 1,301 meters from the swamps. Both commercial and family farms were relatively near to banana trees, including, in some instances, banana growing operations, with an average distance of 85 and 51 meters, respectively. Family and commercial farms were generally close in proximity, with a median distance of 150 meters (IQR 90-300 meters, **Supplemental Figure S5**).

Of the farms that responded (n = 89), 42.7% (n = 38) had piped water, 43.8% (n = 39) primarily got water from a well, while 11.2% (n = 10) primarily used swamp water. The remaining five farms used a combination of rain, swamp, and/or well water. Of those that used well water, almost all reported storing water daily in external containers (n = 38/39), while 26/28, with primarily piped-in water, reported storing water only occasionally.

Out of all farms that reported storing water (n = 85), most water was covered (**Figure 3**), but still 48.7% (37/76) of family farms and 88.9% (8/9) of commercial farms did report having at least some of the water containers as uncovered. The most used container was JerryCans (n = 15, 15, and 5 for those that stored water daily, occasionally, and regularly, respectively).

**Figure 3:**
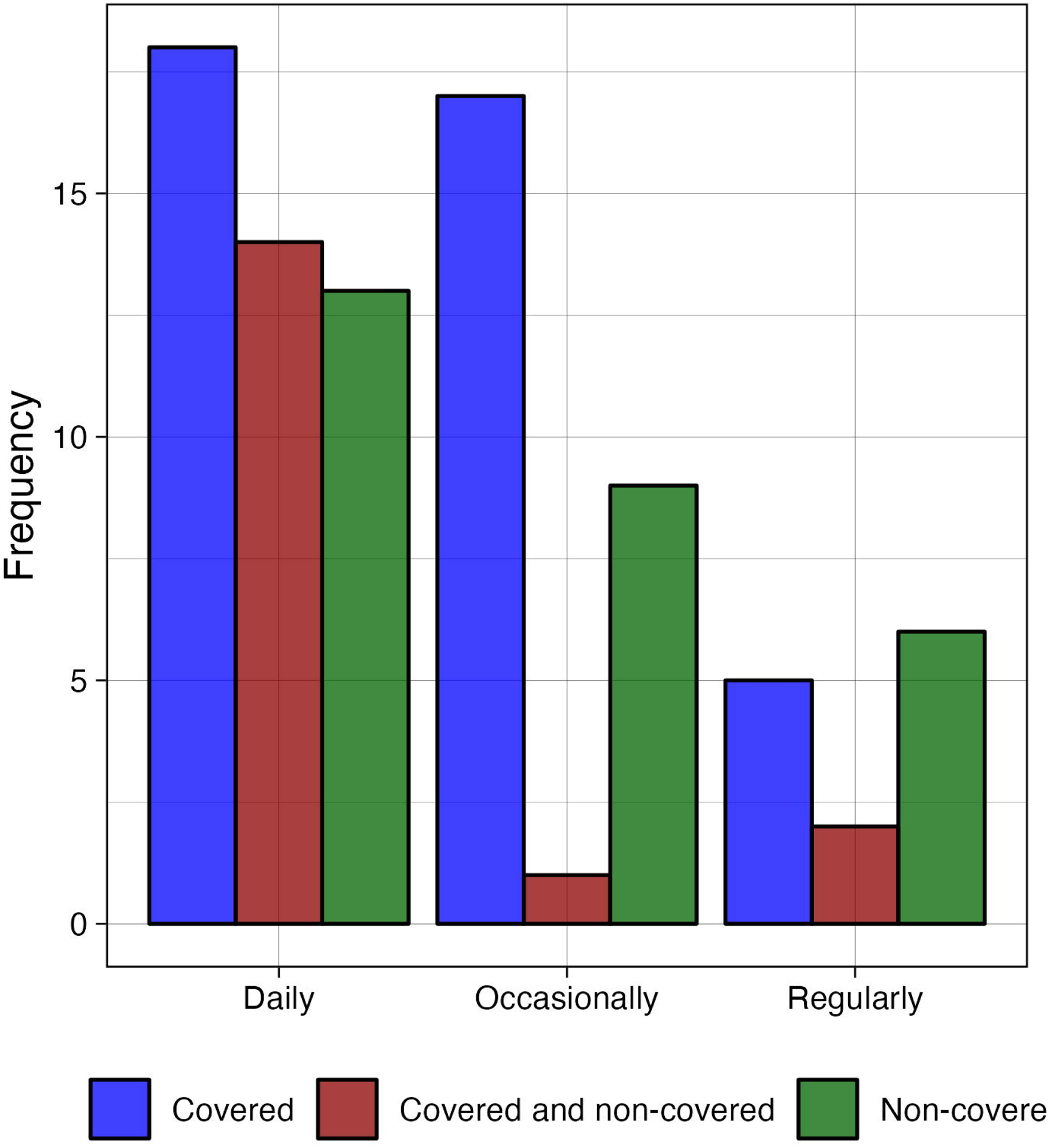
The frequency of whether water was stored (x-axis) and whether that water was kept covered (colored bars).

**Supplemental Table S2** shows all water storage container reporting. At the time of the survey, study personnel noted the presence of water-retaining elements in 45.7% (42/50) of visited farms, 32 (39.02%) of family farms, and 10 (100%) of commercial farms (**Supplemental Figure S6**).

No farms indicated that there was a use of foot bath, regardless of whether family or commercial entity. Further, no farms indicated that new animals were isolated upon acquisition (though 2 commercial and 10 family farms did not provide any information for this question). Five commercial farms (50%) practiced mosquito control within the buildings where humans resided, while 81 of the 82 family farms (98.78%) reported the use of mosquito control inside human dwellings. (Of note: the other six farms provided no information for this question, so it is impossible to determine whether there was no control activity.) When asked about mosquito control specifically for animals, most farms indicated there was none (80% of commercial and 96.34% of family farms) (**Table 4**).

Twenty-seven (20 family and 7 commercial) farms reported a history of abortion in their cattle; 24.4% and 70%, respectively. Only 1 of each farm type reported having repetitive abortions on their farms, while 6 farms (4 commercial and 2 family) did not have record keeping to answer. Of those that reported a history of abortion in cattle, 14 family farms reported that one event had occurred in the last two years, 4 family farms reported two events in the last two years, and 1 family farm reported 3 events in the last two years. Two commercial farms reported 2 events within the last two years, one farm reported 3 events, 1 reported 4 events, 1 reported 6 events, and 1 reported 9 events all within the last two years.

A total of 26 farms (19 family and 7 commercial) reported how abortive waste was disposed of. The most common abortion tissue disposal method was burying without disinfectant (68.4% and 71.4% for family farms and commercial, respectively). Five family farms (26.3%) reported giving abortive tissue to dogs, and one (5.26%) put tissue in normal trash. Two (28.6%) commercial farms gave the abortive tissue to dogs, and no commercial farms disposed in normal trash.

Most farms indicated using bednets; 66 (92.96%) of family farms but only 3 (30%) of commercial farms, though this discrepancy is likely due to the nature of family versus commercial farms and whether personnel sleep at commercial farms. Of those who reported using bed nets, 100% reported using bednets every day. Responders were asked about the history of malaria cases on the farm, and 43 (46.7%) reported no malaria cases since the implementation of bed net use and/or mosquito spraying, while the remaining reported a history of malaria on the farm (**Supplemental Figure S7**). Finally, all but one farm reported disposal of garbage in a large hole, which would eventually be subjected to burning (‘fumier’).

### Environmental Factors as Predictors of Reported Abortion History in Cattle

Ninety farms had a response for whether there was a history of abortion in cattle. Univariate logistic regression was performed with the dependent binary response of abortion history (Yes = 1, No = 0). Altitude, temperature, nor humidity were significantly associated with abortion history (p = 0.393, 0.709, 0.221, respectively). The presence of nearby bushes, swamps, or bananas was also not indicative of a history of abortion (p = 0.5335, 0.1332, 0.149, respectively). Observed elements retaining water by study personnel were also not associated with a history of abortion (p = 0.2147). Cattle source was not significantly associated with abortion history (**Supplemental Tables S3**). Sectors were also not significantly associated with abortion history (**Supplemental Table S4**). Farm type was significantly associated with abortion history (p = 0.00493) with family farms were significantly less likely to have reported abortion history (OR = 0.94).

### Animal Health Factors as Predictors of Reported Abortion History in Cattle

There was no association between the presence or absence of exotic cattle breeds and abortion history (p = 0.5579), the presence or absence of small ruminants (p = 0.56965), or the presence of chickens (p = 0.13924). Species separation in housing practices was not associated with abortion history (p = 0.8826). Zero grazing practices were not associated with abortion history (p = 0.2875). Isolation practices for new animals were also not associated with abortion history in cattle (p = 0.3698), nor were years since the introduction of cattle (p = 0.109146). Further, spearman’s rank correlation revealed that there was no obvious correlation between the number of reported abortion events within the last two years and the total number of cattle on the farm (rho = 0.319, p = 0.1197).

### Human Health Factors as Predictors of Reported Abortion History in Cattle

Distance to human housing from cattle housing was not associated with abortion history (p = 0.201), nor was bed net use (p = 0.6826), nor a history of malaria (p = 0.7821). The water source was not associated with the abortion report, nor was whether water was stored (**Supplemental Tables S5-S6**). The type of storage container (Jerricans vs. other) was not associated with a history of abortion (p=0.2989), nor was coverage of water being stored (**Supplemental Table S7).**

## Associations with the History of Malaria in Households associated with Farms

### Environmental Factors as Predictors of Reported Malaria History in Humans

All 92 farms responded with a history of malaria in humans associated with the farm. Univariate logistic regression was performed with the dependent binary response of malaria history (Yes = 1, No = 0). Sectors were significantly associated with malaria history, with Mpanga differing from the reference group of Gahara. Additionally, when Mpanga was coded as the reference, Kirehe and Nasho were also significantly different from Mpanga. However, when we looked at whether villages (within sectors) were associated with malaria history, there was no effect (**Supplemental Table S8**). Farm type was not significantly associated with malaria history (p = 0.0912). Altitude, temperature, nor humidity were significantly associated with malaria history (p = 0.767, 0.914, 0.912, respectively). The presence of nearby bush or swamp was also not indicative of a history of malaria (p = 0.0757, 0.559, respectively). The presence of bananas near was significantly associated with malaria history (p = 0.026, OR: 3.04) but the distance to banana trees or plantations was not significant (p = 0.209). Observed elements retaining water by study personnel were also associated with a history of malaria (p = 0.0195, OR: 2.76). The type of container that was observed retaining water was not significant (**Supplemental Table S9**).

### Animal Health Factors as Predictors of Reported Malaria History in Humans

There was no association between the presence or absence of exotic cattle breeds and malaria history (p = 0.890), the presence or absence of small ruminants (p = 0.941), or the presence of chickens (p = 0.180). Species separation in housing practices was not associated with malaria history (p = 0.253). Zero grazing practices of cattle were not associated with malaria history (p = 0.604).

### Human Health Factors as Predictors of Reported Malaria History in Humans

Distance to human housing from cattle housing was not associated with malaria history (p=0.634). Water source was not associated with malaria reporting, nor was whether water was stored (**Supplemental Tables S10-S11**). The type of storage container (Jerricans vs. other) was not associated with the history of abortion (p = 0.362), nor was coverage of water being stored (**Supplemental Table S12).** Unsurprisingly, the use of bed nets was significantly associated with reported history of malaria (p = 0.00541), with those who reported the use of bed nets significantly less likely to report a history of malaria (OR: 0.1099). Additionally, reporting the use of bed nets every day was also associated with a decrease in the likelihood of reporting malaria history (p = 0.0074, OR: 0.1183).

Stepwise logistic regression was employed using all significant variables from the univariate analyses and the best model was based on the smallest AIC value over all observations where there were no missing values (n = 84). The final model included the presence of banana trees (y/n), observed elements retaining water (y/n), and the use of bed nets (y/n) (AIC = 106.67). However, the only variable that remained significant was bed net use (yes vs. no, p = 0.0281, OR: 0.1641294).

## Discussion

Animal and environmental health are intimately related to global health. Many leading health organizations have adopted The One Health outlook, including the WHO, FAO, WOAH, and CDC [7, 26, 27]. While not having a uniform definition amongst institutions, One Health is best described as using an integrated and multi-disciplinary approach to investigate and optimize the health of humans, animals, and the environment, and emerged as a popular framework in 2004 in response to the emergence of SARS-1 [28, 29]. This outbreak highlighted the need for a collaborative process that understood that humans, animals, and the environment are closely linked [25, 28, 29]. A One Health approach is particularly beneficial when studying zoonoses and vector-borne pathogens.

In 2021, it was estimated that there were more than 10,000 livestock farms and 430,000 cattle in the Eastern Province of Rwanda [30]. This indicates that cattle and humans are in regular contact. Further, water storage and water accumulation in the environment is a known risk factor for mosquito vector breeding [31, 32]. Because Rwanda experiences regular RVFV outbreaks - its largest in 2018, and because RVFV is a zoonoses in addition to an animal disease of veterinary and economic significance, we focused on the most overt clinical outcome of this virus as an indicator of recognizable risk [14]. These data looked at animal, human, and environmental health factors to determine whether any of these factors could be associated with the risk of zoonotic and/or human mosquito-borne disease. There was no discernable pattern in the significance of risk factors for the history of abortion in cattle associated with these farms from any of the three One Health sectors. However, we were able to identify some potential sources of risk that could be targeted for enhanced education and protection on these farms.

First, mosquito control in the form of source reduction and pesticide use was also problematic. Research shows that cattle can play a positive or negative role in attracting mosquitoes to humans. In cases where cattle live near human sleeping quarters, an increased attraction can be seen, known as zoo-potentiation, and cattle treated with insecticides are associated with a decrease in malaria vectors when used with other vector control approaches [33–35]. Only five farms reported practicing animal mosquito control, and only two identified the product specifically. Two commercial farms reported the use of Norotraz, which is currently only indicated for use by the Rwanda FDA to control ticks [36]. Whether or not this pesticide is efficacious for mosquitoes remains to be widely investigated. The remaining three family farms practiced bush removal as mosquito control. Bushes, especially those with large leaves, can often function as a stagnant water source for mosquito breeding grounds, providing a cover for mosquitoes after breeding [37, 38]. Our study found no significant correlation between the presence and/or distance to the bush and history of either malaria in humans or abortion in cattle. However, a study on integrated malaria management in neighboring Kenya found that removing bushes did reduce the vector population in the home [37]. This study did note a significant relationship between the reported history of malaria and the presence of bananas near the home. The variability in significance and type of surrounding environment may indicate the need for more targeted research on the efficacy of responsible environmental habitat reduction for the control of mosquito vectors.

Second, water storage can be a risk factor for breeding and the presence of mosquito vectors. It is recommended that stagnant water be covered and replaced at least every four days [39]. On average, half of the farms surveyed stored water in their containers daily; though this was not a significant factor, and we do not know the frequency of water turnover in these containers. This study did find a statistically significant association between study personnel observed water accumulation in containers and/or trash in the environment near the home and a history of reported malaria. Bed net use was high in this population, indicating a high level of knowledge regarding the adult mosquito and its risk for transmitting the parasite to the household [40–42]. However, overall, there is an indication that perhaps the juvenile stage of mosquitoes remains unclear to farmers and that there is likely an opportunity for education regarding the mosquito life cycle and the benefits of source reduction for breeding [32]. **Error! Reference source not found.**

Third, the use of biosafety tools, such as footbaths to prevent environmental contamination and the spread of pathogens, was universally lacking in this population, which indicates unawareness of the potential for the spread of this and other cattle-borne viruses and other health conditions in cattle [43–46]. Footbaths can also protect human health, as having a footbath present reduced infection odds by nearly 4-fold [47]. While there was not a significant relationship between abortion history in cattle and tissue disposal methods, we did observe and record risky disposal behaviors. The most common method of tissue disposal in farms with an abortion history was burying without disinfectant, followed by feeding to dogs, and one farm disposed of the tissue in normal trash. Abortive tissue must be disposed of with consideration for the biohazard it represents through any number of methods. Incineration is an effective method for reducing disease transmission but requires specific conditions and likely specialized equipment. Burial is considered the easiest and cheapest option though it invites the potential for ground contamination or disease transmission if not done properly, especially if spore-forming bacteria like *Bacillus anthracis* circulate in the region [48]. Further, Rift Valley Fever and other bunyaviruses have been shown to be environmentally stable, which increases the risk of environmental or fomite-mediated transmissions [49–51]. Personal protective equipment, such as gloves and masks, and other preventative measures should be taken when handling abortive tissue.

Our study is not without limitations. As a cross-sectional study of attitudes and practices, it is difficult to determine historical effectors of transmission and risk. Further, this survey was conducted before the COVID-19 global pandemic, which, at best, resulted in an increase in public awareness of infection control. A follow-up study would be necessary to determine whether there have been any changes in the use of PPE or infection control methods on farms, given the recency of the pandemic. Another limitation of our study appears in the relatively limited sample size of commercial farms, which may have affected our ability to make comparisons, with only ten commercial farms relative to eighty-two family farms. However, our data does provide a helpful building block to begin assessing the social and environmental determinants of One Health assessments on Rwandan farms.

## Conclusion

Considering our findings, policymakers and health officials should focus on promoting personal protective equipment (PPE) and preventative measures on livestock farms. Emphasizing the importance of safe abortive tissue disposal, proper mosquito control, and preventive practices like foot baths for cattle can significantly improve animal health and mitigate disease risks. Addressing these issues can positively affect livestock farms, safeguarding animal welfare and farmers’ livelihoods. The WHO continually speaks of the importance of education on vector-borne diseases and promoting behavior change [52]. This study showcases the health of rural Rwandan farms and highlights areas of improvement for animal and human health.

## Data Availability

All data produced in the present work are contained in the manuscript

## Acknowledgements

We would like to thank Jean Claude Tumushime, Evodie Uwimbabazi, Elisee Ndizeye, Grace Mukasine, Jean Bosco Noheri, Jean de Dieu Ayabagabo, and Christopher Mores for their assistance and guidance. As always, thank you to Autumn for her unwavering attitude and to Julie, Alex, Isabella, Alexandra, and Liliana Espino for their support.

## Disclaimer

The contents are solely the responsibility of the authors and do not necessarily represent the official views of the USDA or NIFA.

