## Supplemental Information for "Assessment of risk factors associated with and practices of cattle farmers in Kirehe District Rwanda with respect to vector-borne and zoonotic pathogens"

**Supplemental Information**
Dutuze, Espino, and Christofferson.

**Supplemental Figure S1:** Altitude (A), temperature (B), and humidity (C) measures for each sector within the Kirehe District where farms were sampled.

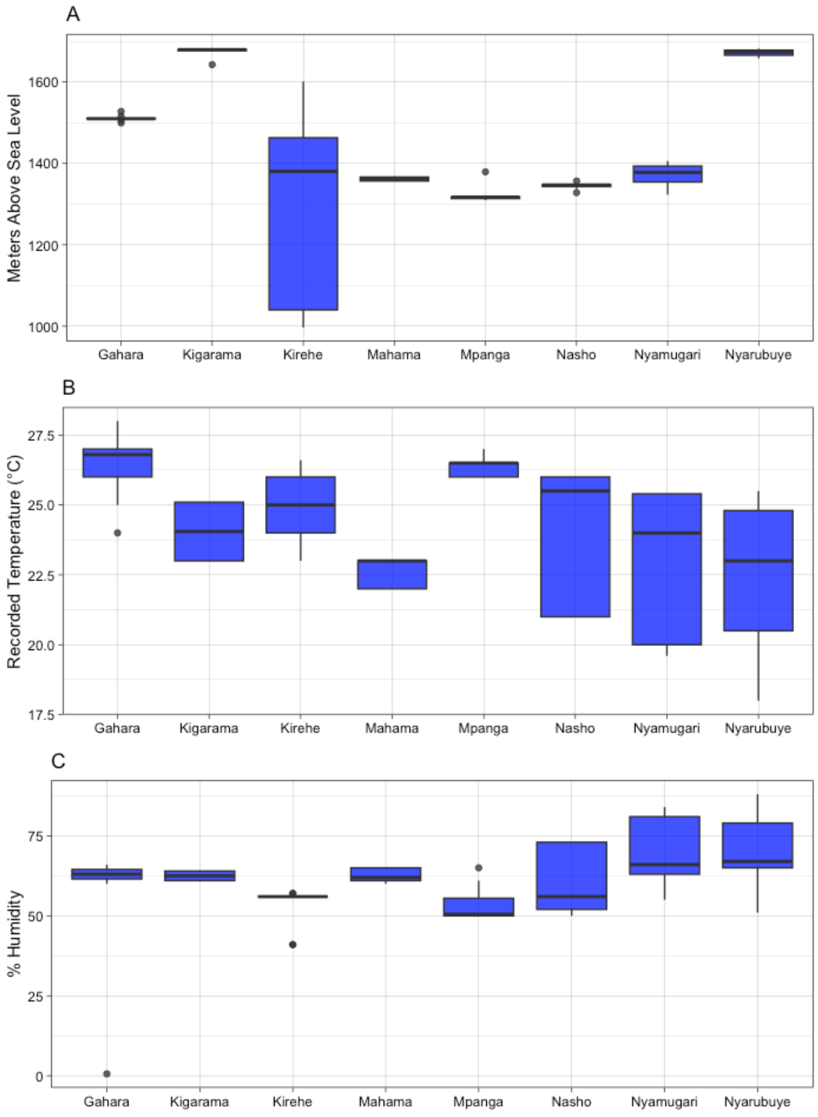

**Supplemental Figure S2:** Linear fit between altitude and humidity and temperature. Temperature and humidity were significantly, but moderately associated with altitude (adjusted $R^2$ of 0.04628 and .1077, respectively).

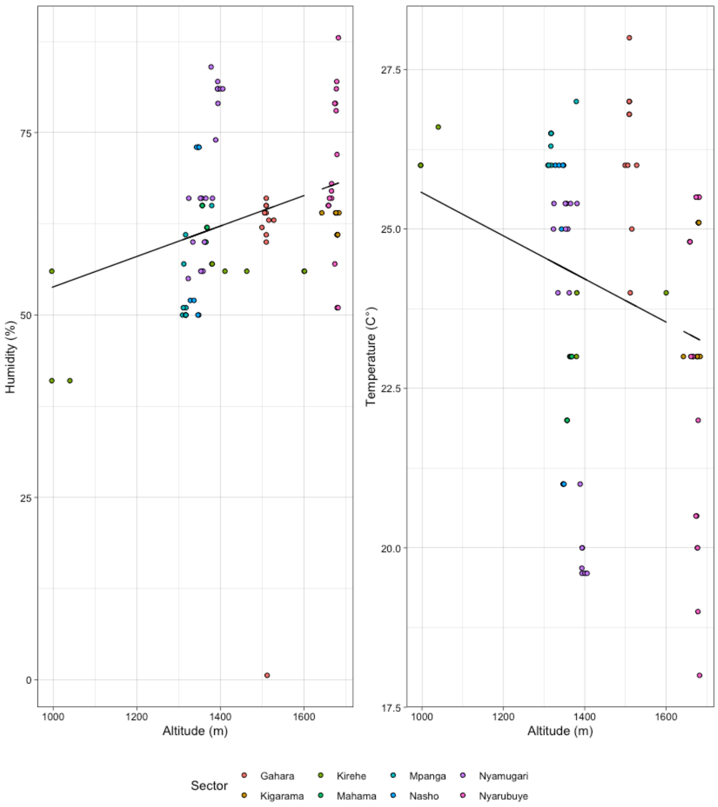


**Supplemental Table S1:** The exotic and local breed numbers of cows for the 89 farms that reporting housing cows and reported the number of cattle on the farm.

| Farm Type | Number of Farms with Exotic cow breed | Average number of exotic cow breed | Number of Farms with Local cow breed | Average number of local cow breed | Average total number of cows |
|---|---|---|---|---|---|
| Commercial | 9 | 20 | 9 | 5.12 | 25.8 |
| Family | 72 | 0.627 | 72 | 2.39 | 2.93 |

**Supplemental Figure S3**: Box plot of the distance to bush recorded for commercial (n=10) and family (n=82) farms sampled as measured by study personnel depending on the response by the responder as to whether they considered themselves to be nearby to the bush (Yes vs. No).

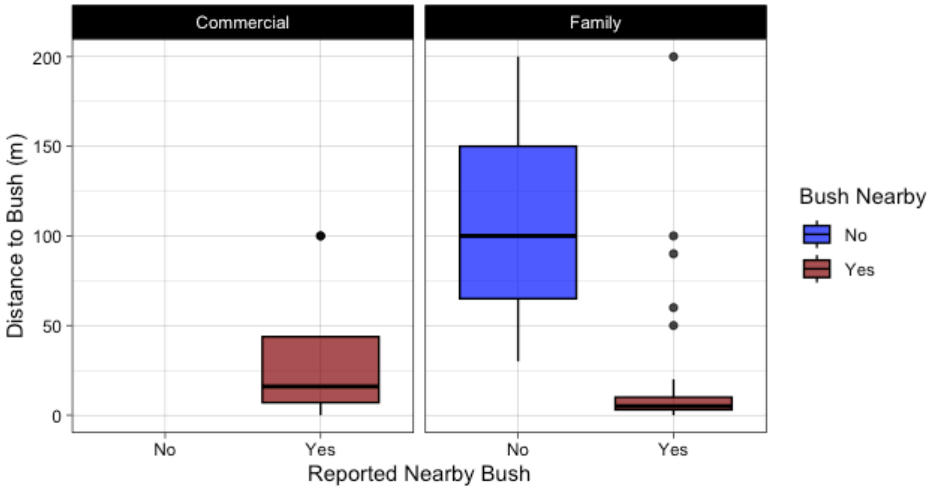

**Supplemental Figure S4**: Box plot of the distance to swamp recorded for commercial (n=10) and family (n=82) farms sampled as measured by study personnel depending on the response by the responder as to whether they considered themselves to be nearby to swamp (Yes vs. No).

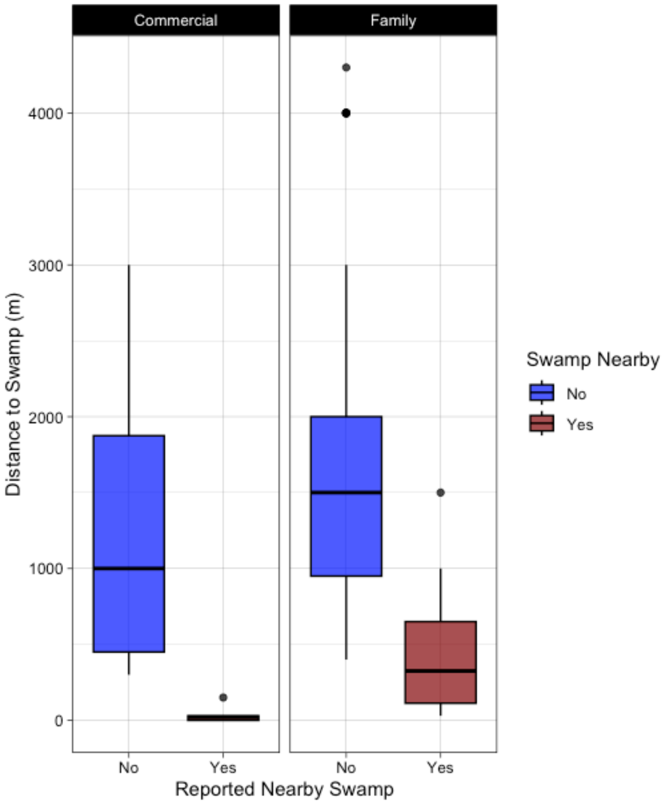

**Supplemental Figure S5**: Boxplot of distance to nearest commercial farms for all family farms (left) and by Sector (right).

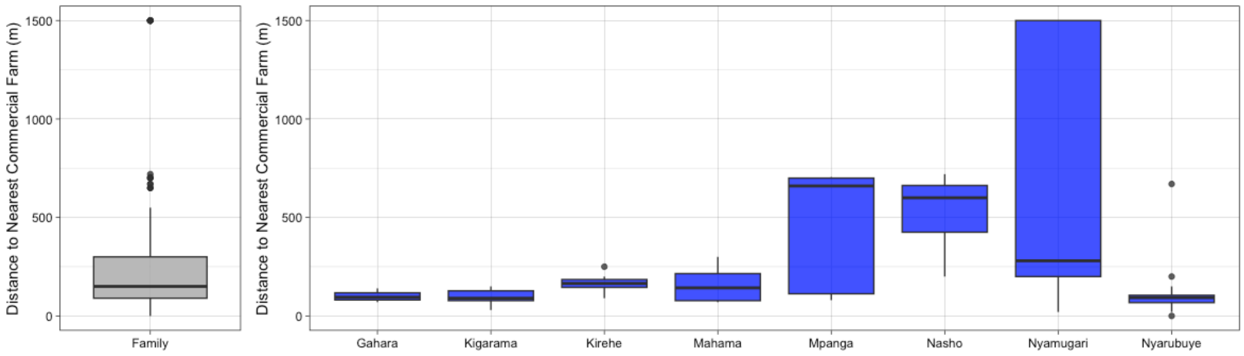


**Supplemental Table S2**: Water Storage habits of the 85 farms that reporting storing water (9 commercial and 76 family farms) either daily, occasionally, or regularly.

| | Daily | Occasionally | Regularly |
|---|---|---|---|
| Backets only | 4 | 1 | 0 |
| Backets & JerryCans | 2 | 0 | 0 |
| Bassines | 1 | 0 | 1 |
| Clays | 0 | 0 | 1 |
| Drinkers | 0 | 0 | 1 |
| JerryCans | 14 | 15 | 5 |
| JerryCan pieces | 0 | 1 | 0 |
| JerryCans & Sheeting | 1 | 0 | 0 |
| JerryCans & Tanks | 9 | 1 | 2 |
| Sheetings | 6 | 1 | 0 |
| Tanks | 7 | 0 | 3 |
| Tanks & Drinkers | 1 | 9 | 0 |

**Supplemental Figure S6**: Observed containers or other materials containing water recorded by study personnel. (Imivure: containers made from wood; Ibibumbiro: containers made from concrete, usually for providing water to animals; Ibibindi: containers made from clay.)

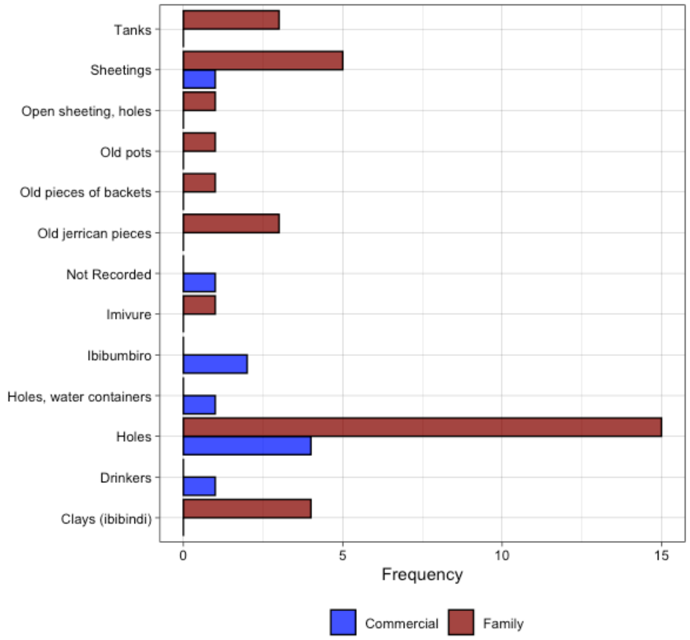


**Supplemental Figure S7**: The number of farms reporting a history of malaria during the months specified. Six (6) farms did not provide information on when malaria cases occurred.

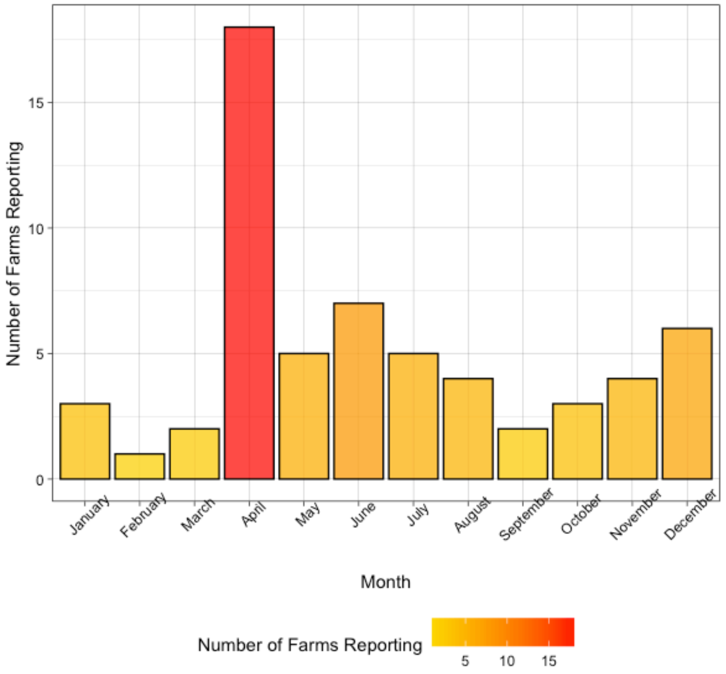

**Supplemental Table S3**: Estimates and p-values associated with logistic regression for abortion history according to source of cattle. Estimate is log(odds).

| | Estimate | p-value |
|---|---|---|
| Family (reference level) | --- | --- |
| Government | -0.34 | 0.64 |
| Market | 0.07 | 0.91 |
| Multiple Sources | -1.10 | 0.36 |
| Neighbor | -15.87 | 0.99 |

**Supplemental S4**: Estimates and p-values associated with logistic regression for abortion history according to sector. Estimate is log(odds).

| | Estimate | p-value |
|---|---|---|
| Gahana (reference level) | --- | --- |
| Kigarama | -2.01 | 0.10 |
| Kirehe | -1.07 | 0.29 |
| Mahama | -0.51 | 0.63 |
| Mpanga | -0.22 | 0.80 |
| Nasho | -1.76 | 0.15 |
| Nyamugari | -0.85 | 0.29 |
| Nyarubuye | -0.17 | 0.82 |

**Supplemental Table S5:** Estimates and p-values associated with logistic regression for abortion history according to water source. Estimate is log(odds).

| | Estimate | p-value |
|---|---|---|
| Piped (reference level) | --- | --- |
| Swamp | 1.46 | 0.06 |
| Swamp, rain | 0.76 | 0.56 |
| Well | 0.76 | 0.16 |
| Well, swamp | 17.02 | 0.99 |

**Supplemental Table S6:** Estimates and p-values associated with logistic regression for abortion history according to frequency of storage of water near the farm. Estimate is log(odds).

| | Estimate | p-value |
|---|---|---|
| Daily (reference level) | --- | --- |
| None | -0.15 | -0.17 |
| Occasionally | -0.67 | -1.13 |
| Regularly | 0.61 | 0.95 |

**Supplemental Table S7:** Estimates and p-values associated with logistic regression for abortion history according to whether stored water was covered. Estimate is log(odds).

| | Estimate | p-value |
|---|---|---|
| Covered (reference level) | --- | --- |
| Mixed | 0.22 | 0.73 |
| Non-covered | 0.63 | 0.25 |

**Supplemental Table S8:** Estimates and p-values associated with logistic regression for malaria history according to sector. Estimate is log(odds).

| | Model 1 | | Model 2 | |
|---|---|---|---|---|
| | Estimate | p-value | Estimate | p-value |
| Gahara (reference level for Model 1) | --- | --- | 2.37 | 0.02* |
| Kigarama | -0.98 | 0.29 | 1.39 | 0.17 |
| Kirehe | -0.29 | 0.77 | 2.08 | 0.05* |
| Mahama | -1.90 | 0.08 | 0.47 | 0.68 |
| Mpanga (reference level for Model 2) | -2.37 | 0.02* | --- | --- |
| Nasho | 0.27 | 0.80 | 2.64 | 0.02* |
| Nyamugari | -0.88 | 0.29 | 1.49 | 0.10 |
| Nyarubuye | -0.86 | 0.30 | 1.50 | 0.11 |

**Supplemental Table S9:** Estimates and p-values associated with logistic regression for malaria history according to the types of containers study personnel observed retaining water. Estimate is log(odds). (Imivure: containers made from wood; Ibibumbiro: containers made from concrete, usually for providing water to animals; Ibibindi: containers made from clay.)

| | Estimate | p-value |
|---|---|---|
| Clays/ Ibibindi (reference level) | --- | --- |
| Drinkers | -17.57 | 1.00 |
| Holes | 0.54 | 0.63 |
| Holes, water containers | 17.57 | 1.00 |
| Ibibumbiro | 2.88 e-15 | 1.00 |
| Imivure | 17.57 | 1.00 |
| NA | -0.34 | 0.75 |
| Not Recorded | 17.57 | 1.00 |
| Old jerrican pieces | 0.69 | 0.66 |
| Old pieces of backets | -17.57 | 1.00 |
| Old pots | 17.57 | 1.00 |
| Open sheeting, holes | 17.57 | 1.00 |
| Sheetings | 1.61 | 0.28 |
| Tanks | 0.69 | 0.66 |

**Supplemental Table S10:** Estimates and p-values associated with logistic regression for malaria history according to source of water. Estimate is log(odds).

| | Estimate | p-value |
|---|---|---|
| Piped (reference level) | --- | --- |
| Rain | 15.78 | 0.99 |
| Swamp | 0.62 | 0.39 |
| Swamp, rain | 0.90 | 0.48 |
| Well | 0.47 | 0.31 |
| Well, swamp | 15.78 | 0.99 |

**Supplemental Table S11:** Estimates and p-values associated with logistic regression for malaria history according to frequency of storage of water near the farm. Estimate is log(odds).

| | Estimate | p-value |
|---|---|---|
| Daily (reference level) | --- | --- |
| No | -1.42 | 0.11 |
| Occasionally | -0.42 | 0.39 |
| Regularly | -0.97 | 0.14 |

**Supplemental Table S12:** Estimates and p-values associated with logistic regression for malaria history according to whether stored water was covered. Estimate is log(odds).

| | Estimate | p-value |
|---|---|---|
| Covered (reference level) | --- | --- |
| Mixed | 0.56 | 0.34 |
| Non-covered | 0.95 | 0.07 |
